# Single-cell multi-omics analysis reveals IFN-driven alterations in T lymphocytes and natural killer cells in systemic lupus erythematosus

**DOI:** 10.1101/2021.04.27.21256106

**Authors:** Dominik Trzupek, Mercede Lee, Fiona Hamey, Linda S. Wicker, John A. Todd, Ricardo C. Ferreira

**Affiliations:** JDRF/Wellcome Diabetes and Inflammation Laboratory, Wellcome Centre for Human Genetics, Nuffield Department of Medicine, NIHR Oxford Biomedical Research Centre, University of Oxford, Oxford, UK

**Keywords:** Single-cell RNA-sequencing (scRNA-seq), multi-omics, BD Rhapsody AbSeq, cytotoxic CD4^+^ T cells (CTLs), systemic lupus erythematosus (SLE), type I interferon (IFN), biomarker

## Abstract

**Background:** The characterisation of the peripheral immune system in the autoimmune disease systemic lupus erythematosus (SLE) at the single-cell level has been limited by the reduced sensitivity of current whole-transcriptomic technologies. Here we employ a targeted single-cell multi-omics approach, combining protein and mRNA quantification, to generate a high-resolution map of the T lymphocyte and natural killer (NK) cell populations in blood from SLE patients.

**Methods:** We designed a custom panel to quantify the transcription of 534 genes in parallel with the expression of 51 surface protein targets using the BD Rhapsody AbSeq single-cell system. We applied this technology to profile 20,656 T and NK cells isolated from peripheral blood from an SLE patient with a type I interferon (IFN)-induced gene expression signature (IFN^hi^), and an age- and sex- matched IFN^low^ SLE patient and healthy donor.

**Results:** We confirmed the presence of a rare cytotoxic CD4^+^ T cell (CTL) subset, which was exclusively present in the IFN^hi^ patient. Furthermore, we identified additional alterations consistent with increased immune activation in this patient, most notably a shift towards terminally differentiated CD57^+^ CD8^+^ T cell and CD16^+^ NK^dim^ phenotypes, and the presence of a subset of recently-activated naïve CD4^+^ T cells.

**Conclusions:** Our results identify IFN-driven changes in the composition and phenotype of T and NK cells that are consistent with a systemic immune activation within the IFN^hi^ patient, and underscore the added resolving power of this multi-omics approach to identify rare immune subsets. Consequently, we were able to find evidence for novel cellular peripheral biomarkers of SLE disease activity, including a subpopulation of CD57^+^ CD4^+^ CTLs.

## Introduction

Systemic lupus erythematosus (SLE) is a common autoimmune disease, characterised by chronic immune activation including a systemic type I interferon (IFN)-induced transcriptional signature in blood (henceforth referred to as the IFN signature), and the development of anti-nucleic acid autoantibodies^1,2^. The immunological complexity of SLE is underscored by the extensive heterogeneity of the clinical manifestations, which impair the management of the condition and is reflected in the paucity of tailored therapeutic options available to the patients. In particular, the presence of the IFN signature has been shown to be associated with increased disease severity^3,4^. These findings have been widely replicated and establish the pathogenic role of the IFN signalling pathway in SLE^5,6^. Furthermore, the detection of an IFN signature in blood can be used to stratify patients with chronic immune activation, and several surrogate biomarkers- for example soluble SIGLEC-1 (sSIGLEC-1) and IL-2 concentrations and a population of CD25^−^FOXP3^+^ Tregs - have been proposed to be useful to identify patients with active disease^7–9^. However, the exact mechanisms underlying the pathological chronic immune activation are only partly identified, particularly the contribution of the adaptive immune system to this condition. The recent advent of high-throughput single-cell RNA-sequencing (scRNA-seq) has provided a new tool to dissect the extensive heterogeneity of the human immune system and to identify disease-specific alterations. Nevertheless, these methods still present significant technical limitations, which include – but are not limited to – high cost and low sensitivity, especially for genes with low expression levels, that can significantly reduce the power of the analyses. In SLE, a recent scRNA-seq study did demonstrate the applicability of the approach to dissect the heterogeneity of the immune response at the single-cell level^10^. Nevertheless, the limited sensitivity of the whole-transcriptome method precluded the identification of disease-specific alterations in the T cell compartment over-and-above the previously identified IFN-induced gene expression signature.

Recently, we have developed a targeted single-cell multi-omics approach based on the BD Rhapsody system combined with the AbSeq proteomics technology, which allows us to immunophenotype the human immune system with high resolution and in a large number of cells in a more affordable way^11^. This approach, combining mRNA and protein quantification provides superior sensitivity and resolution compared to traditional whole-transcriptomic methods, which are critical factors for the single-cell characterisation of relatively quiescent cells with low RNA content such as T cells. Using this approach, we have identified a rare subset of activated Th1 CD4^+^ T cells with a distinct cytotoxic signature in blood from an SLE patient with a high IFN-induced gene expression signature (IFN^hi^)^11^. To replicate and extend these findings, we here applied this technology to three additional selected blood samples: (i) an IFN^hi^ SLE patient with concomitant high circulating IL-2 levels; (ii) an age- and sex-matched IFN^low^ SLE patient; and (iii) an age- and sex-matched healthy donor. To further increase the sensitivity of this system to dissect the heterogeneity of T and NK cell populations, we designed a custom mRNA panel containing 565 different probes targeting 534 different genes, including a selection of genes with high cell-type discrimination potential identified in other T and NK cell datasets, thereby providing increased resolution for the clustering of these immune populations. In addition, we developed a panel of 51 AbSeq antibodies, targeting key T- and NK-cell enriched surface markers, which provide additional functional insight into the identified cell subsets. This resource provides a high-resolution map of the peripheral T and NK populations in SLE, containing ∼21,000 high-quality profiled single cells, and provides a proof-of-principle framework for the application of this multi-omics approach to elucidate the cellular mechanisms contributing to autoimmune diseases.

## Methods

### Ethics statement

All samples and information were collected with written and signed informed consent from all participants. The study conformed to the principles of the Helsinki Declaration and was approved by the NHS Peterborough and Fenland local research ethics committee (05/Q0106/20).

#### Subjects

Study participants comprised two SLE patients and one healthy donor, recruited from the Cambridge BioResource (CBR). These three donors were selected from a cohort of 490 healthy donors and 41 SLE patients from the CBR, which were previously recruited to explore novel biomarkers of the IFN signature and disease activity in SLE^8,9^. Based on the blood biomarkers previously measured in this extended cohort, we selected three patients to profile in this study: an IFN^hi^ SLE patient with concomitant high circulating IL-2 levels, one age-and sex-matched IFN^low^ SLE patient and one age- and sex-matched healthy donor. Baseline demographics and characteristics are summarised in Table 1. The SLE patients were recruited for this study outside their regular clinic visits and represent population-based patients with good disease management that were otherwise well at the time of venesection.

**Table 1.**
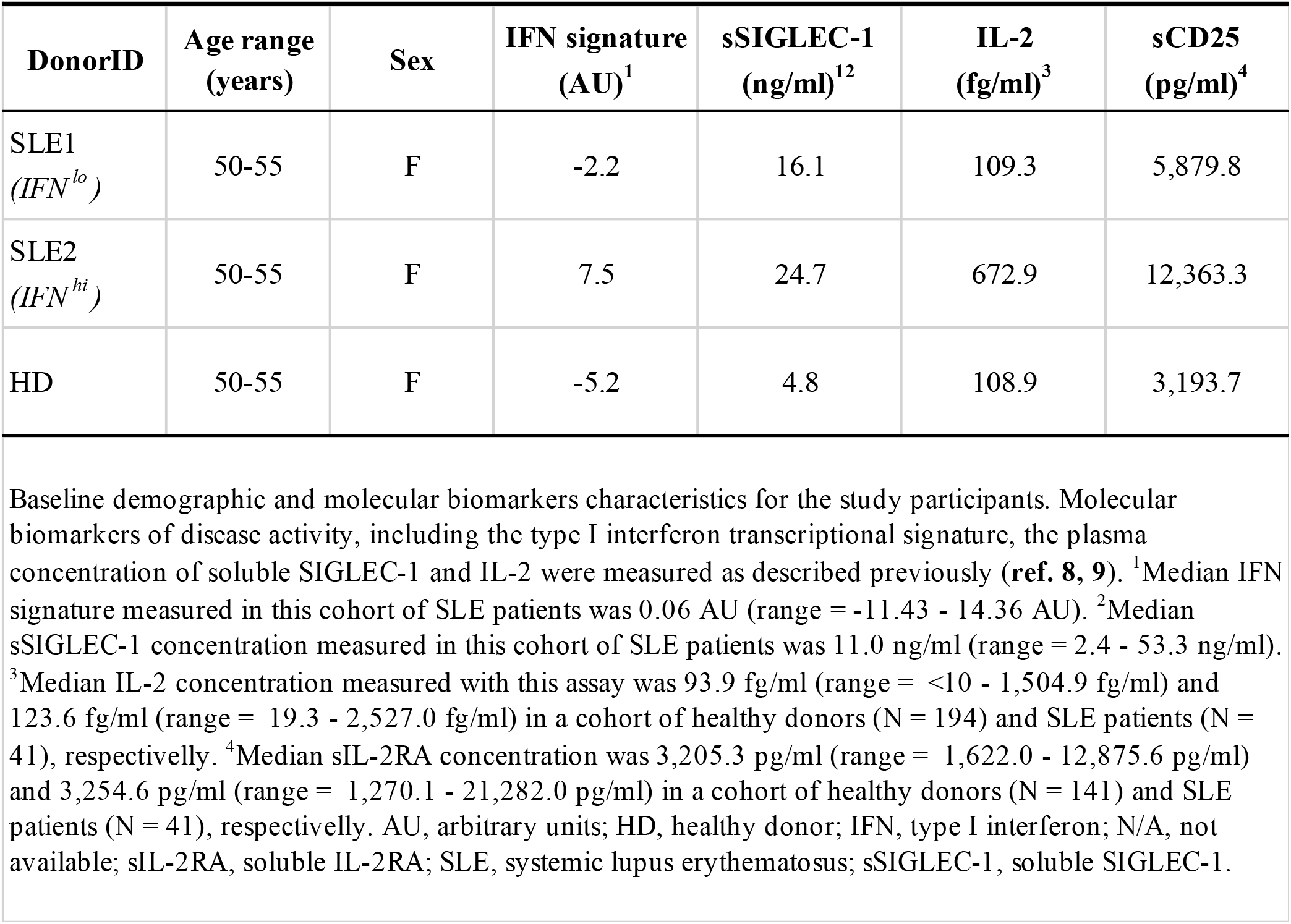
Baseline characteristics of study participants.

| DonorID | Age range (years) | Sex | IFN signature (AU)<sup>1</sup> | sSIGLEC-1 (ng/ml)<sup>12</sup> | IL-2 (fg/ml)<sup>3</sup> | sCD25 (pg/ml)<sup>4</sup> |
| --- | --- | --- | --- | --- | --- | --- |
| SLE1<br>(*IFN<sup>lo</sup>*) | 50-55 | F | -2.2 | 16.1 | 109.3 | 5,879.8 |
| SLE2<br>(*IFN<sup>hi</sup>*) | 50-55 | F | 7.5 | 24.7 | 672.9 | 12,363.3 |
| HD | 50-55 | F | -5.2 | 4.8 | 108.9 | 3,193.7 |

Baseline demographic and molecular biomarkers characteristics for the study participants. Molecular biomarkers of disease activity, including the type I interferon transcriptional signature, the plasma concentration of soluble SIGLEC-1 and IL-2 were measured as described previously (**ref. 8, 9**). <sup>1</sup>Median IFN signature measured in this cohort of SLE patients was 0.06 AU (range = -11.43 - 14.36 AU). <sup>2</sup>Median sSIGLEC-1 concentration measured in this cohort of SLE patients was 11.0 ng/ml (range = 2.4 - 53.3 ng/ml). <sup>3</sup>Median IL-2 concentration measured with this assay was 93.9 fg/ml (range = <10 - 1,504.9 fg/ml) and 123.6 fg/ml (range = 19.3 - 2,527.0 fg/ml) in a cohort of healthy donors (N = 194) and SLE patients (N = 41), respectively. <sup>4</sup>Median sIL-2RA concentration was 3,205.3 pg/ml (range = 1,622.0 - 12,875.6 pg/ml) and 3,254.6 pg/ml (range = 1,270.1 - 21,282.0 pg/ml) in a cohort of healthy donors (N = 141) and SLE patients (N = 41), respectively. AU, arbitrary units; HD, healthy donor; IFN, type I interferon; N/A, not available; sIL-2RA, soluble IL-2RA; SLE, systemic lupus erythematosus; sSIGLEC-1, soluble SIGLEC-1.

#### Cell preparation and fluorescence-activated cell sorting (FACS)

Cryopreserved peripheral blood mononuclear cells (PBMCs) were thawed at 37°C and resuspended drop-by-drop in X-VIVO15 (Lonza) with 1% heat-inactivated, filtered human AB serum (Sigma). After washing in PBS, cells were incubated with Fixable Viability Dye eFluor 780 (eBioscience) for 10 minutes at room temperature. Cells were then washed with PBS + 2% FBS and incubated with human Fc block (BD Biosciences) and with the following fluorochrome conjugated antibodies: CD25-PE (Mouse anti-human monoclonal antibody, clone M-A521; BD Biosciences, cat# 555432), CD127-PE cy7 (Mouse anti-human monoclonal antibody, clone eBioRDR5; eBioscience; cat# 25-1278-42), CD4-FITC (Mouse anti-human monoclonal antibody, clone OKT4; Biolegend, cat# 317408), CD56-BV711 (Mouse anti-human monoclonal antibody, clone HCD56; Biolegend, cat# 318336), CD8-AF700 (Mouse anti-human monoclonal antibody, clone HIT8a; Biolegend, cat# 300920) and CD3-BV510 (Mouse anti-human monoclonal antibody, clone UCHT1; Biolegend, cat# 300448). Following incubation for five minutes, each sample was split in two 5ml FACS tubes (Falcon) and labelled with the respective sample barcoding antibody (sample multiplexing kit; BD Biosciences). Each donor was barcoded with two barcodes (six barcodes in total) to increase the number of sample barcodes and improve the identification of cell multiplets. The two barcodes corresponding to the same donor were collapsed into a single sample at the analysis stage. Following incubation for 30 minutes at room temperature, cells were washed two times and resuspended in PBS + 1% FBS for cell sorting at 4°C in a BD FACSAria Fusion sorter (BD Biosciences). The following T and NK cell subsets were sorted from each donor: (i) CD3^−^CD56^hi^ (NK56^br^); (ii) CD3^−^CD56^+^ (NK56^dim^); (iii) CD3^+^CD8^+^ (CD8^+^ T cells); (iv) CD3^+^CD4^+^ CD127^hi^CD25^−/low^ (CD4^+^ CD127^hi^ Teffs); (v) CD3^+^CD4^+^ CD127^low^CD25^−/low^ (CD4^+^ CD127^low^ Teffs); and (vi) CD3^+^CD4^+^ CD127^low^CD25^hi^ (CD4^+^ Tregs).

#### Plasma soluble IL-2RA (sCD25) measurements

Soluble IL-2RA (sCD25) concentrations were measured in plasma samples from a cohort of SLE patients (N=41) and healthy donors (N=141), which included the three donors profiled in this study, using the BD OptEIA Human ELISA Kit (BD Biosciences; cat# 559104) according to the manufacturer’s instructions. Assay readout was performed using Europium-labelled streptavidin (Perkin Elmer, cat# 1244-360) combined with time-resolved fluorescence spectroscopy using DELFIA reagents (DELFIA Assay buffer, cat# 1244-106 and DELFIA Enhancement Solution, cat# 1244-104, Perkin Elmer). Plasma samples were diluted 1:20 and measured in duplicate in Corning 96-well clear polystyrene microtiter plates (Sigma, cat# CL3795-100EA) using a VICTOR X4 multilabel plate reader (Perkin Elmer). Each 96-well plate contained a recombinant sCD25 protein standard curve with a detection range of 31.25– 500pg/ml.

#### AbSeq staining and cell capture

Following cell sorting, each cell subset was resuspended to a final concentration of 200,000 cells/ml. To capture similar numbers of cells from each subset and each donor, 100 µl of each subset (corresponding to 20,000 cells) from the same donor were then pooled together. For the NK56^br^ subset, cell numbers obtained from sorting were limited (ranging from 4,300 to 10,000), and therefore all cells were taken to the final donor pool. Cells were then washed and resuspended in 300 µl X-VIVO15 + 10% heat-inactivated, filtered human AB serum. Half of the volume from each donor (corresponding to ∼55,000 cells) was pooled together in a 5 ml FACS tube and washed with PBS + 2% FBS. Cells were stained with a master mix of 51 oligo-conjugated AbSeq antibodies (BD Bioscience; *see extended data*) for 45 minutes on ice.

The other half of the volume from each donor was incubated in round-bottom 96-well plates at 37°C for 90 minutes with a PMA and ionomycin cell stimulation cocktail (eBioscience), in the absence of protein transport inhibitors. Cells were harvested into FACS tubes, washed with PBS + 2% FBS and incubated with the oligo-conjugated AbSeq antibodies, as detailed above for the resting cells.

Following incubation with AbSeq antibodies, cells were washed three times with cold BD sample buffer (BD Biosciences) to remove any unbound antibody and counted. Samples were then resuspended in 620 µl of cold BD sample buffer at the desired cell concentrations – ranging from 20 to 30 cells/µl for an estimated capture rate of 10,000-15,000 single-cells – and immediately loaded on a BD Rhapsody cartridge (BD Biosciences, cat# 633733; and cartridge reagent kit, BD biosciences, cat# 633731) for single-cell capture. Resting and *in vitro* stimulated samples were each loaded on a separate BD Rhapsody cartridge.

#### cDNA library preparation and sequencing

Single-cell capture and cDNA library preparation was performed using the BD Rhapsody Express Single-cell analysis system (BD Biosciences), using the BD Rhapsody cDNA (BD Biosciences, cat# 633663) and Targeted amplification (BD Biosciences, cat# 633664) kits, according to the manufacturer’s instructions. cDNA was amplified for 11 cycles using the pre-designed Human T-cell Expression primer panel (BD Biosciences, cat# 633751) containing 259 primer pairs, together with a custom designed primer panel containing 306 primer pairs (BD Biosciences, cat# 633784). Our combined panel contained 565 primer pairs targeting 534 different genes (see *extended data*). Panel design was based on previous literature search combined with data mining of available T and NK cell datasets to identify a panel of highly variable genes that contribute highly to the clustering of the identified T and NK cell subsets. This panel was specifically enriched for lineage-defining transcription factors and other key functional markers of Th cell differentiation to increase clustering resolution.

The resulting PCR1 products were purified using AMPure XP magnetic beads (Beckman Coulter, cat#A63881) and the respective mRNA and AbSeq/Sample tag products were separated based on size selection, using different bead ratios (0.7X and 1.2X, respectively). The purified mRNA and sample tag PCR1 products were further amplified (10 cycles), and the resulting PCR2 products purified by size selection (0.8X and 1.2X for the mRNA and sample tag libraries, respectively). The concentration, size and integrity of the resulting PCR products was assessed using both Qubit (High Sensitivity dsDNA kit; Thermo Fisher, cat# Q32854) and the Agilent 4200 Tapestation system (High Sensitivity D1000 screentape; Agilent, cat#5067-5584 and High sensitivity D1000 Reagents; Agilent, cat# 5067-5585). The final products were normalised to 2.5 ng/µl (mRNA), 1.1 ng/µl (Sample tag) and 0.5 ng/µl (AbSeq) and underwent a final round of amplification (six cycles) using indexes for Illumina sequencing to prepare the final libraries. Final libraries were quantified using Qubit and Agilent Tapestation and pooled (∼39/58/3% mRNA/AbSeq/Sample tag ratio) to achieve a final concentration of 5 nM. Final pooled libraries were spiked with 15% PhiX control DNA to increase sequence complexity and sequenced (75 bp paired-end) on a HiSeq 4000 sequencer (Illumina).

#### Data analysis and quality control (QC)

The FASTQ files obtained from sequencing were analysed following the BD Biosciences Rhapsody pipeline (BD Biosciences), as previously described^11^, using as alignment reference the pre-designed Human T-cell expression panel containing 259 amplicon sequences (mapping to 259 different genes; BD Biosciences), a custom supplemental panel containing 306 amplicon sequences (mapping to an additional 275 genes), and an AbSeq panel of 51 oligo sequences. Reads with the same cell label, same unique molecular identifier (UMI) sequence and mapping to the same gene were collapsed into a single molecule. The recursive substitution error correction (RSEC) UMI adjustment algorithm developed by BD Biosciences was used to obtain the adjusted UMI counts for further processing and analysis.

A total of 10,973 (resting) and 14,509 (stimulated) putative cells were called by the Rhapsody data processing pipeline, of which 583 and 1,427, respectively, corresponded to pseudo-cells containing only very low mRNA and protein counts – due to low-level ambient mRNA contamination. From the 10,390 (resting) and 13,082 (stimulated) captured cells, 321 (3.9%) and 547 (4.2%), respectively, were assigned as cell multiplets based on their sample barcode information and filtered out from the analysis. Additionally, we excluded cells with highest (>40,000) UMI counts. Most cells identified as undetermined by the Rhapsody pipeline could be confirmed to be of low quality apart from the subset of NK56^br^ cells identified as undetermined due to low affinity of antibodies used for multiplexing towards these cells. All remaining undetermined cells not mapping to the identified NK56^br^ cluster were excluded from the analysis. Additionally, small subsets of cells of clearly distinct clusters were removed due to being identified as either apoptotic cells (characterised by lower distribution of mRNA UMI counts and higher expression of all AbSeq antibodies – due to higher levels of unspecific antibody binding on apoptotic cells), or non-T cell contaminants (small numbers of B cells and monocyte contaminants). From the initial 10,390 (resting) and 13,082 (stimulated) captured cells, we ended up with a final set of 9,568 and 11,088 cells, respectively, after QC, corresponding to a final processed dataset of 20,656 high-quality single-cells. All processed scRNA-seq data files containing the gene expression matrixes (UMI per cell) and sample tag identifiers for the resting, stimulated and integrated datasets are available as underlying data (Dataset S1)^12^.

Data normalisation was performed using different methods for the protein and mRNA data: centred log-ratio (CLR) normalisation for protein, computed independently for each feature; and typical log normalisation for the mRNA data. We used Uniform Manifold Approximation and Projection (UMAP) for dimensionality reduction, increasing the number of PCA dimensions to 30, based on the Seurat elbow plot. Differential expression analysis was performed using a tailored hurdle model from the MAST package^13^, and integration of data from multiple experiments was performed using a combination of canonical correlation analysis (CCA) and identification of mutual nearest neighbours (MNN), implemented in Seurat 3.0^14^. For clustering of the resting dataset, we increased the default clustering resolution parameter value to 1.1 to obtain a more fine-grained set of clusters. Although we observed relatively stable cluster assignment around this value of the clustering parameter, the optimal parameter may depend on the aim of the analysis and on the desired granularity of the resulting cell clusters.

## Results

### Single-cell multi-omics provides a high-resolution map of T and NK cell populations in blood

Previously, we have demonstrated that a targeted single-cell multi-omics approach, based on the BD Rhapsody system (using a pan-immune panel containing 399 probes), provides a high-resolution single-cell map of the human peripheral immune system using PBMCs^11^. To further refine the resolution of the T and NK cell populations, we have developed a custom targeted mRNA panel containing 565 primer pairs targeting 534 genes, with an emphasis on key lowly-expressed, lineage-defining, transcripts. Of the 565 primers included in this custom panel, only a subset of 260 overlapped with the pan-immune panel we used previously^11^, thereby providing increased resolution to identify the functional T and NK cell subsets. We have applied this custom panel, in combination with 51 oligo-conjugated AbSeq antibodies, to profile the T and NK cell populations in PBMCs in SLE (Figure 1A). We were interested in investigating alterations in these immune compartments in SLE patients with a recent IFN response, so we specifically selected one independent IFN^hi^ patient sample in which we had also previously measured elevated sSIGLEC-1, IL-2 and sCD25 concentrations, as well an age- and sex-matched IFN^low^ patient and a healthy donor^8,9^. To reduce experimental variation, we used oligo-conjugated barcode antibodies to identify each of the three donors and pool the cells into a single experiment (Figure 1A). Furthermore, to increase the number of T and NK cells profilled and therefore increase the ability to detect new subpopulations of cells and biomarkers, we employed a FACS-sorting strategy to specifically enrich for six immune subsets of interest: (i) CD3^−^ CD56^hi^ NK cells (NK56^br^); (ii) CD3^−^ CD56^+^ NK cells (NK56^dim^); (iii) CD3^+^ CD4^−^CD8^+^ T cells (CD8^+^ T cells); (iv) CD3^+^ CD4^+^CD8^−^ CD127^hi^CD25^-/low^ T cells (CD4^+^ CD127^hi^ Teffs); (v) CD3^+^ CD4^+^CD8^−^ CD127^low^CD25^−/low^ T cells (CD4^+^ CD127^low^ Teffs); and (vi) CD3^+^ CD4^+^CD8^−^ CD127^low^CD25^hi^ T cells (CD4^+^ Tregs; Figure 1B). To obtain a similar representation of each of these immune subsets we aimed to pool equal numbers (∼20,000) of cells sorted from each subset, with the exception of the more limiting NK56^br^ subset where we aimed to sort ∼10,000 cells per donor. We noted a significant depletion of NK56^br^ cells in the IFN^hi^ patient (sorted 4,300 cells – compared to 10,000 for the IFN^low^ patient and healthy donor), which may indicate a mobilization of the circulating NK56^br^ cells to tissues because of the IFN response in this patient (Figure 1C).

**Figure 1.**
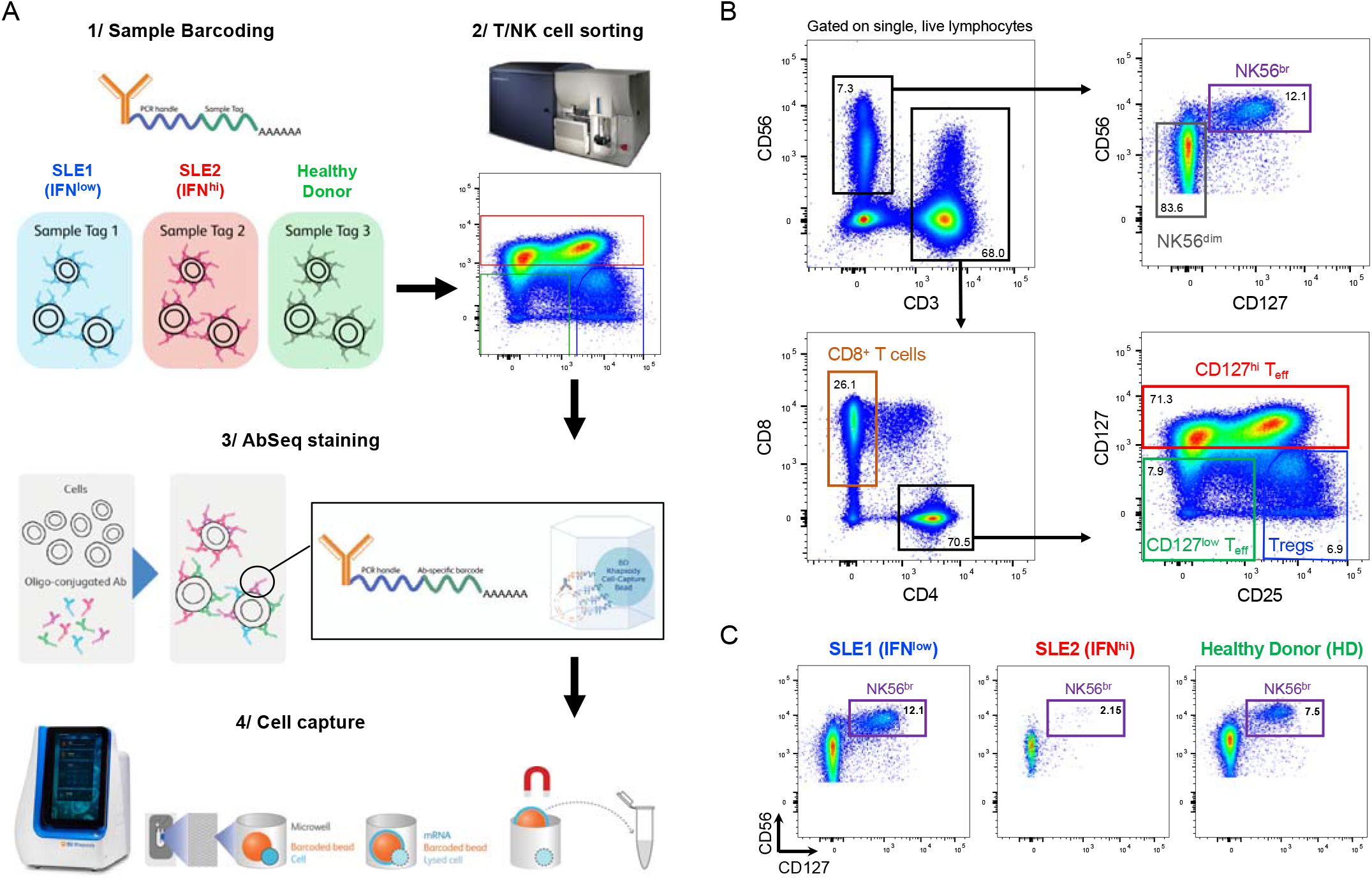
Characterisation of the peripheral T and NK cell populations in SLE using a single-cell multi-omics approach. (**A**) Summary of the experimental workflow based on the BD Rhapsody single-cell RNA-sequencing system combining the quantification of mRNA and surface protein targets (AbSeq). Experiment was carried out using samples from three donors: (i) an SLE patient with low expression of the type I interferon (IFN) transcriptional signature (IFN^low^; depicted in blue); (ii) an SLE patient with a previously detected IFN signature (IFN^hi^; depicted in red); and (iii) and healthy donor (HD; depicted in green). Cells isolated from each donor were barcoded with oligo-conjugated antibodies and pooled together for cell capture and library preparation. (**B**) Gating strategy used for the isolation of the six T and NK cell populations assessed in this study. (**C**) Frequency of NK56^br^ cells, defined as CD3^-^CD127^+^CD56^hi^ NK cells, in each of the three donors.

To obtain a more comprehensive map of the assessed subsets, we obtained single-cell transcriptomics and proteomic profiles from both resting and *in vitro* stimulated cells. In total we profiled 20,656 single cells with high quality, corresponding to 9,568 resting and 11,088 *in vitro* stimulated cells. For the resting cells, unsupervised clustering identified 15 distinct subsets, with a clear demarcation of cells from the main CD4^+^, CD8^+^ and NK cell populations (Figure 2A, 2B; underlying data – Dataset S2)^12^. This improved functional differentiation of the T cell compartment is illustrated by the mutually exclusive pattern of expression of the CD45RA and CD45RO protein isoforms, reflecting the transition of a naïve to a memory phenotype (Figure 2C). Furthermore, expression of the CD45RA/RO isoforms on the CD8^+^ T cells identifies a subset of effector memory CD8^+^ T cells within cluster 4 marked by the re-expression of CD45RA, which is compatible with a terminally differentiated effector memory T cell (TEMRA) phenotype. Although we observed good integration of the data from the three donors in this experiment, we observed some marked alterations within the relative distribution of IFN^hi^ patient subsets, manifested in the donor-specific enrichment of four of the identified subsets: two CD4^+^ T cell subsets (clusters 10 and 12), the CD8^+^ Tem/TEMRA subset (cluster 4) and one CD16^+^ NK^dim^ (cluster 3) subset (Figure 2D, 2E). These observations support the presence of subtle yet defined IFN-induced alterations in the composition of rare immune subsets that can be identified using this targeted single-cell multi-omics approach.

**Figure 2.**
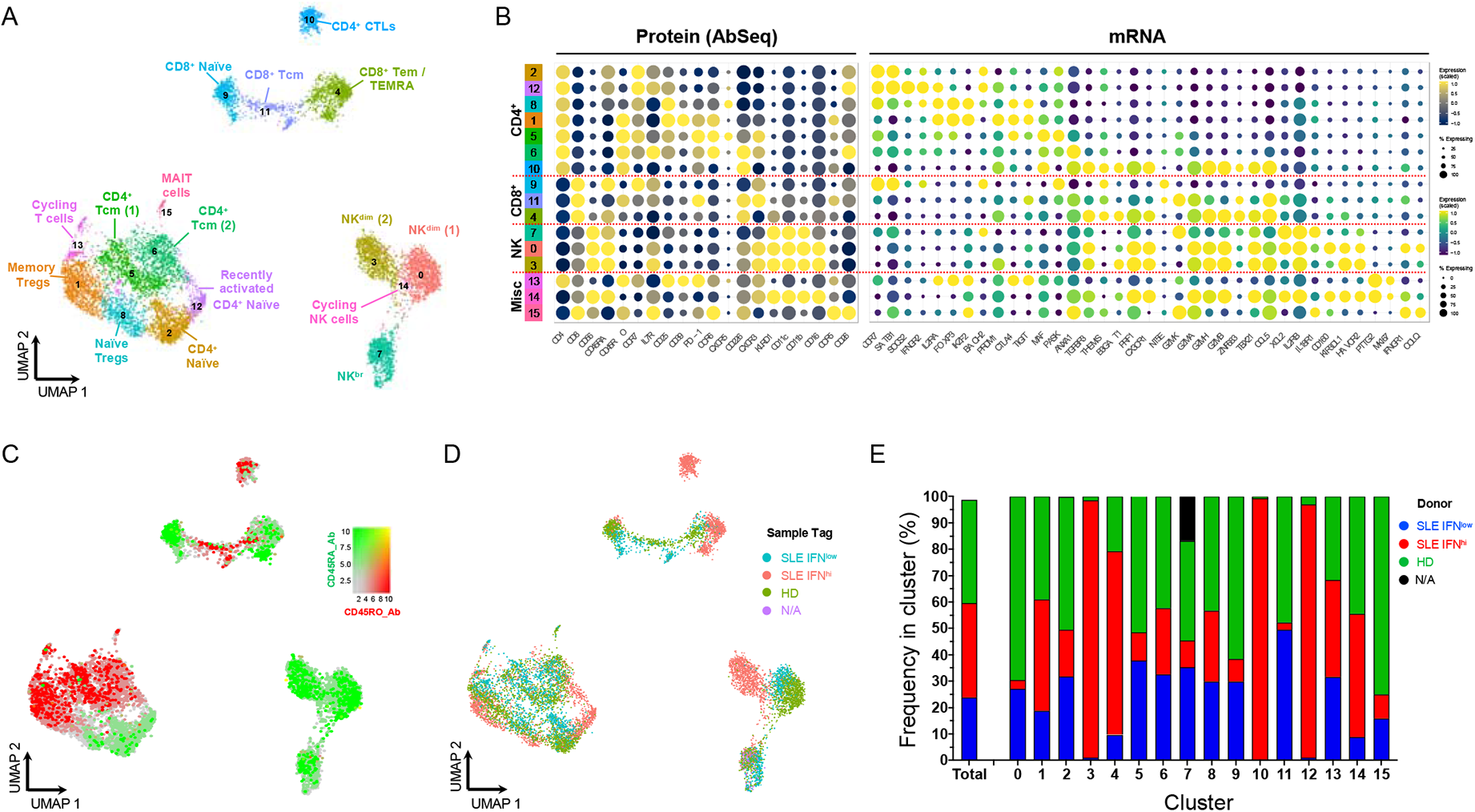
Single-cell multi-omics provides a high-resolution map of the peripheral T and NK cell populations. (**A**) Uniform Manifold Approximation and Projection (UMAP) plot depicting the clustering of the isolated resting cells (N = 9,568 cells passing QC) from the three donors assessed in this study. (**B**) Dot plot depicts the expression of select key differentially expressed markers within the identified clusters. The size of the dots represents the frequency of the marker in the respective cluster and the colour represents the relative expression level. Protein markers (AbSeq) are shown on the left section of the plots and mRNA markers on the right section. (**C**) UMAP plot depicting the overlaid expression of the CD45RA (white to green) and CD45RO (white to red) protein isoforms on the identified cell clusters. (**D**) UMAP plot depicting the donor-specific clustering. (**E**) Compositional analysis depicting the relative frequency of cells from each donor in each of the identified clusters. A small fraction of the NK56^br^ cells in cluster 7 could not be tagged with the barcoding antibodies, and therefore donor-specific identity could not be assigned for those cells (labeled as N/A on the plot). HD, healthy donor; IFN, type I interferon transcriptional signature; SLE, systemic lupus erythematosus.

### A subset of CD4^+^ T cells with cytotoxic profile is specifically present in blood from IFN^hi^ >patients

Among the four IFN^hi^-enriched clusters identified in this study, one corresponded to a CD4^+^ T cell subset (cluster 10) with a clearly-defined cytotoxic profile. Because of this cytotoxic transcriptional profile this subset clustered together with the effector memory CD8^+^ T cell subsets, despite its elevated CD4 expression and concomitant lack of CD8 expression observed by AbSeq (Figure 3A). Differential expression analysis confirmed a distinct cytotoxic profile in comparison with other CD4^+^ T cells, including elevated expression of canonical T cell cytotoxic markers such as PRF1, GZMB, NKG7 and CCL5 (Figure 3B, 3C; underlying data – Dataset S3)^12^. Notably, we had previously identified a CD4^+^ T cell subset with a similar cytotoxic profile using this targeted multi-omics approach that was also specifically enriched in a different IFN^hi^ SLE patient^11^. The T-cell centric custom panel used in this study provides additional resolution of this subset and lends further support to its specific enrichment in IFN^hi^ patients. Of the differentially expressed markers that compose the cytotoxic signature associated with this CD4^+^ CTL subset, we identified the expression of a surface protein, CD57 (encoded by *B3GAT1*; Figure 3D). Similarly, additional genes encoding for surface proteins typically expressed on cytotoxic CD8^+^ T cells, but not in CD4^+^ T cells, including CX3CR1 and KLRG1, were specifically upregulated in these CD4^+^ CTLs (Figure 3D). In addition to cell surface markers, CD57^+^ CD4^+^ CTLs were characterised by the expression of key transcription factors associated with T cell cytotoxic function, including *ZNF683* (Hobit), *IKZF1* (IKAROS), *TBX21* (T-Bet), *ZEB2, RUNX3, ID2, PRDM1* (BLIMP1) and *BHLHE40* (Figure 3B; underlying data – Dataset S3)^12^. These observations suggest that a combination of the CD4 and CD57 markers could be sufficient to identify this subset of circulating CD4^+^CD57^+^ CTLs, and that the frequency of these cells could therefore represent a novel cellular biomarker of recent, and probably flaring, disease activity in SLE patients.

**Figure 3.**
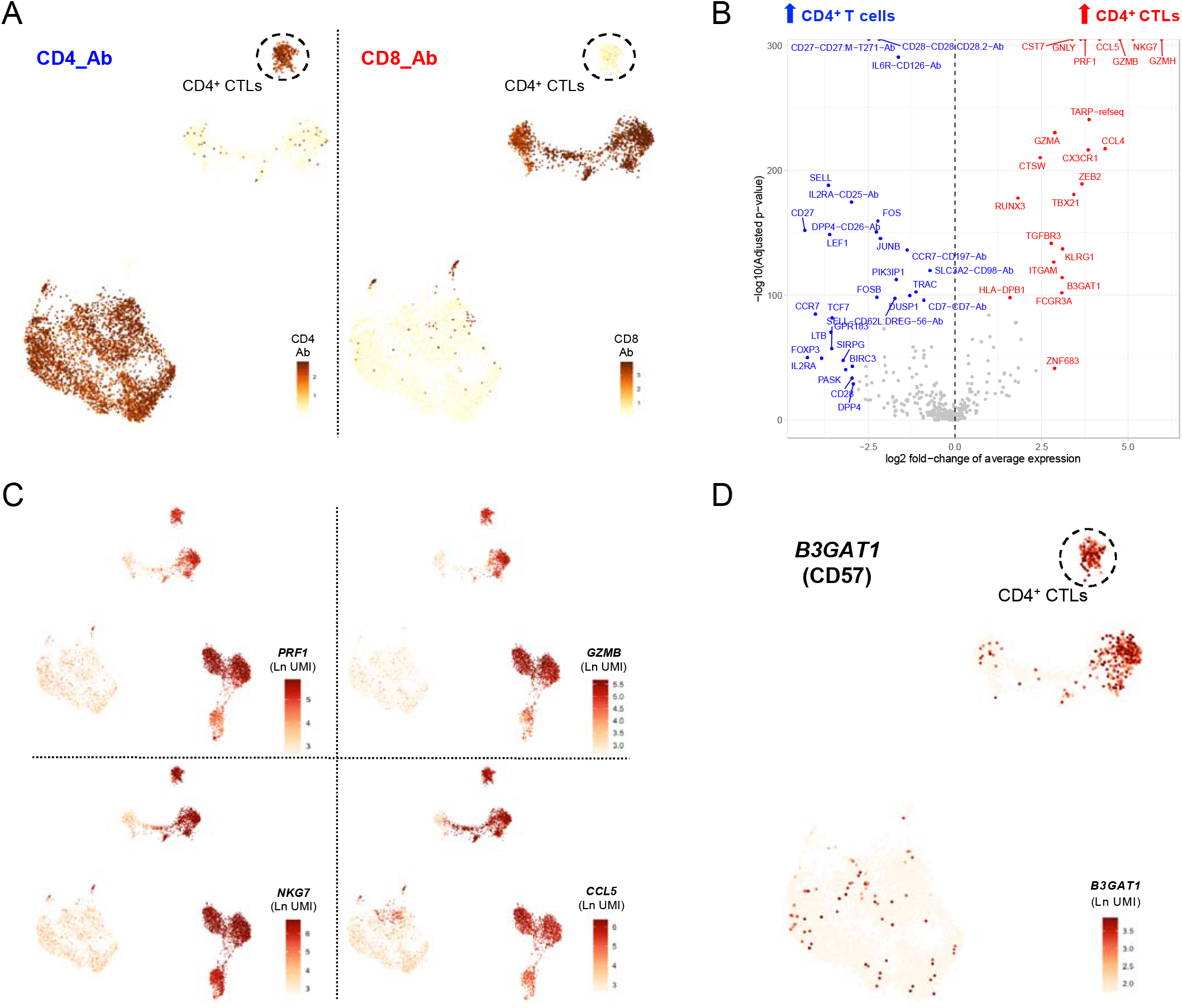
A subset of CD57^+^ CD4^+^ T cells with cytotoxic profile is specifically enriched in the IFN^hi^ SLE patient. (**A**) Feature plots depicting the expression of the CD4 (left) and CD8 (right) at the protein level (AbSeq) on the identified T cell clusters. (**B**) Volcano plot depicts the differential expression between the CD4^+^ CTLs (cluster 10) and the remaining identified CD4^+^ T cell clusters. (**C**) Feature plots depict the expression of four canonical cytotoxic cytokine genes: *PRF1, GZMB, NKG7* and *CCL5*. (**D**) Feature plot depicting the expression of *B3GAT1*, encoding for the surface receptor CD57 on the identified T cell clusters. Ab, AbSeq antibody; CD4^+^ CTL, cytotoxic CD4^+^ T cell.

### In vitro stimulation polarises the functional differentiation of T cells

To further increase the resolution of the functional T cell subsets identified with this multi-omics approach, we also profiled cells following a short (90 minutes) *in vitro* stimulation with PMA + ionomycin. The induction of cytokine expression and upregulation of effector function genes increased further the separation of the subpopulations (Figure 4A, 4B; underlying data – Dataset S4)^12^. This functional separation was particularly evident in CD4^+^ T cells, where we were able to identify the different Th subsets, marked by the expression of the respective lineage-defining transcription factors and canonical effector cytokines (Figure 4C). We were also able to integrate both the resting and *in vitro* stimulated datasets, thereby providing increased cell numbers from each of the identified subsets (Figure 5A, 5B; underlying data – Dataset S5)^12^. The resulting clusters from the integrated analysis generally contained an informative representation of cells from both resting and *in vitro* stimulated conditions, with only a few condition-specific clusters remaining - mostly resulting from the strong induction of cytokine and effector gene expression following *in vitro* stimulation (Figure 5C). In agreement with the efficient integration of the two datasets, the compositional analysis of the integrated dataset yielded similar results to the resting cells (Figure 5D). Only a few exceptions were detected, including the split between the resting and *in vitro* stimulated CD4^+^ CTL clusters (clusters 14 and 17, respectively), which was due to the differential expression of increased levels of Th1-like pro-inflammatory cytokines, such as IFN-*γ*, upon stimulation (Figure 5B). Nevertheless, both clusters of CD4^+^ CTLs were almost exclusively detected within the IFN^hi^ SLE patient (Figure 5D). In addition to this alteration in the CD4^+^ T cell compartment, we also observed a notable shift in the relative composition of the CD8^+^ T cell population within the IFN^hi^ SLE patient, with a strong reduction in the proportion of naïve CD45RA^+^ CD8^+^ T cells (cluster 5) and a concomitant increase in the proportion of terminally differentiated CD8^+^ Tem/TEMRA (cluster 11) cells (Figure 5E).

**Figure 4.**
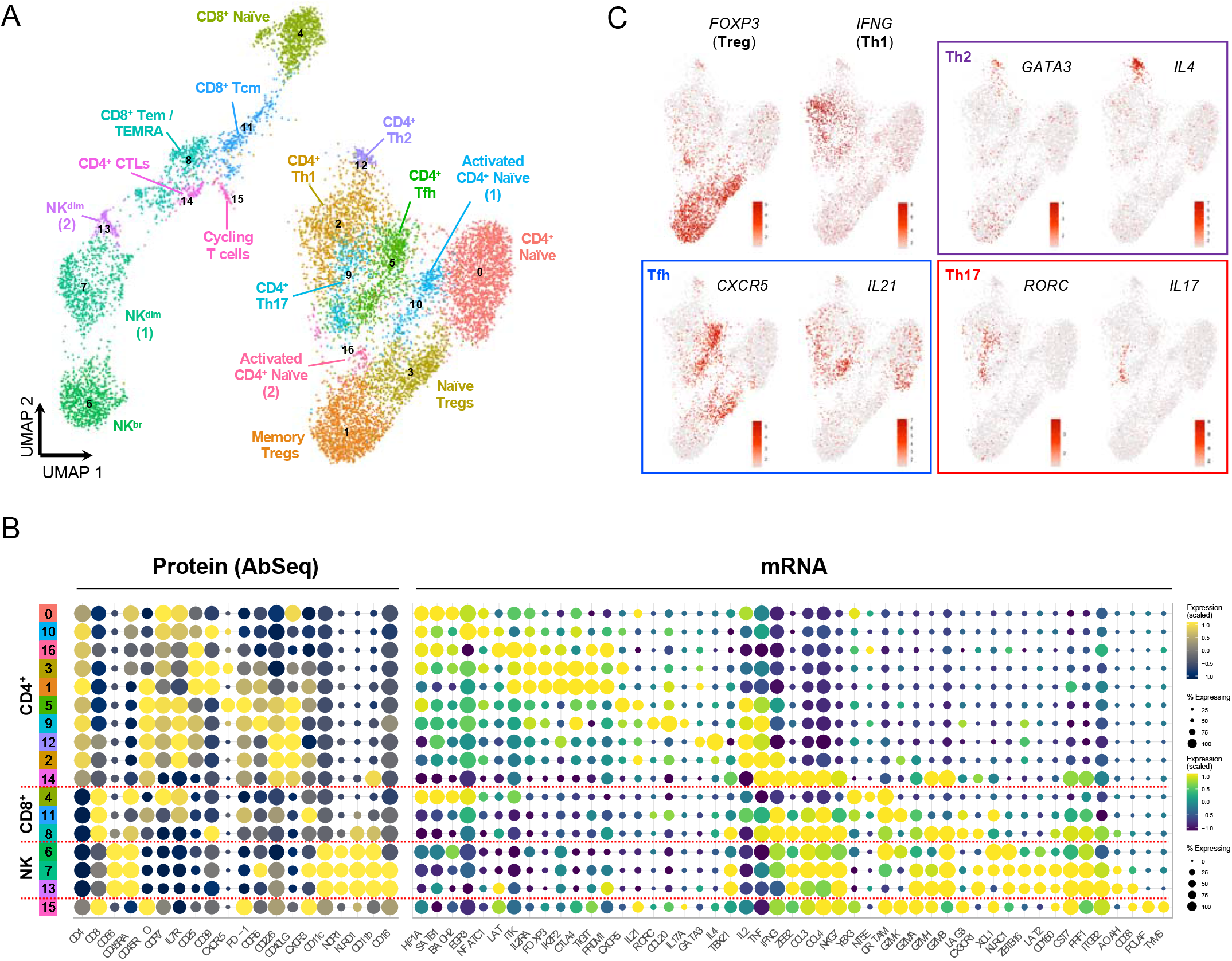
*In vitro* stimulation polarizes the functional differentiation of CD4^+^ T cell subsets. (**A**) UMAP plot depicting the clustering of the isolated T and NK cells following short *in vitro* stimulation (90 min) with PMA + ionomycin (N = 11,088 cells passing QC). (**B**) Dot plot depicts the expression of select key differentially expressed markers within the identified *in vitro* stimulated clusters. The size of the dots represents the frequency of the marker in the respective cluster and the colour represents the relative expression level. Protein markers (AbSeq) are shown in the left section of the plots and mRNA markers on the right section. (**C**) Feature plots depicting the expression of key lineage defining transcription factors and respective effector cytokine.

**Figure 5.**
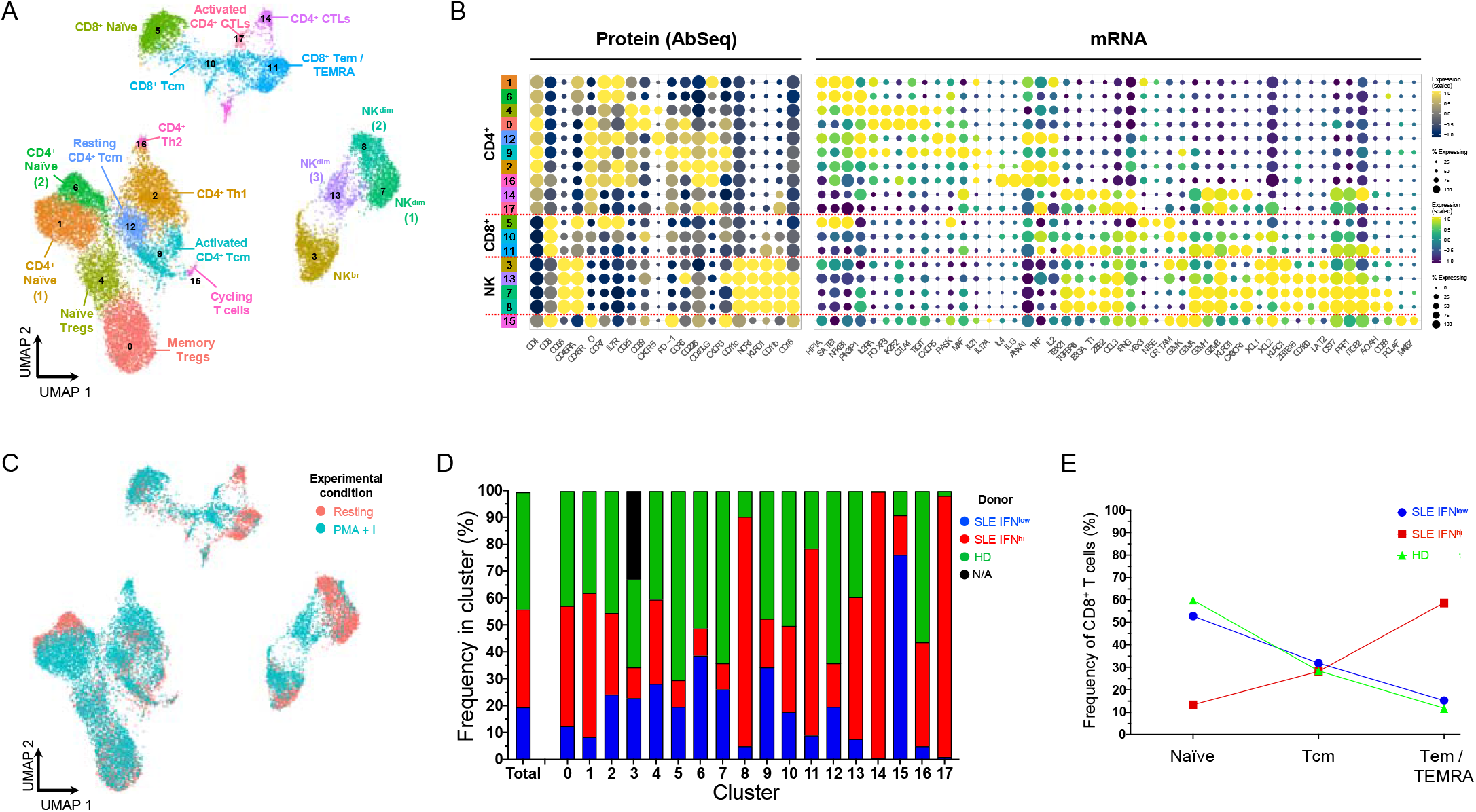
Integrated resting and *in vitro* stimulated data reveal subtle IFN-induced alterations in the composition of the peripheral T and NK cell populations. (**A**) UMAP plot depicting the clustering of integrated (resting + *in vitro* stimulated) T and NK datasets (N = 20,656 cells passing QC). (**B**) Dot plot depicts the expression of select key differentially expressed markers within the identified integrated clusters. The size of the dots represents the frequency of the marker in the respective cluster and the colour represents the relative expression level. Protein markers (AbSeq) are shown on the left section of the plots and mRNA markers on the right section. (**C**) UMAP plot depicts the treatment-specific clustering of the resting (red) and *in vitro* stimulated (teal) cells. (**D**) Compositional analysis depicting the relative frequency of cells from each donor in each of the identified clusters. A small fraction of the NK56^br^ cells in cluster 3 could not be tagged with the barcoding antibodies, and therefore donor-specific identity could not be assigned for those cells (labeled as N/A on the plot). (**E**) Relative distribution of the three identified CD8^+^ T cell subsets (among total CD8^+^ T cells) in each of the three donors assessed in this study. HD, healthy donor; IFN, type I interferon transcriptional signature; SLE, systemic lupus erythematosus; Tcm, central memory T cell; Tem, effector memory T cell; TEMRA, terminally differentiated effector memory T cell.

### CD16^+^ NK^dim^ cells from the IFN^hi^ patient display an activated profile

In addition to the T cell compartment, NK cells isolated from the three donors clustered in four subsets: one cluster of CD16^−^ NK56^br^ cells and three clusters of CD16^+^ NK^dim^ cells. Using the combined transcriptomics and proteomics profile of these cells, we were able to classify the identified NK cell clusters according to a gradient of activation, reflected by the acquisition of an increasingly cytolytic function - from the least cytotoxic NK56^br^ cells (cluster 3) to the more activated CD16^+^ NK^dim^ cells (clusters 7 and 8; Figure 6A, based on NK cell clusters shown in Figure 5A). Among the conventional CD16^+^ NK^dim^ cells, we identified a separation into two subsets (clusters 7 and 8), which was driven by a gradient of expression of markers usually expressed in terminally-differentiated cells, such as *LAG3* and *B3GAT1* (CD57), and the transcription factor *PRDM1* (encoding BLIMP-1) in cluster 8 (Figure 6B, 6C; underlying data – Dataset S6)^12^. We observed marked donor-specific differences in the relative distribution of these NK^dim^ subsets, with the more differentiated cluster 8 being almost exclusively detected in the IFN^hi^ patient (Figure 5D). These data are consistent with the highly differentiated CD8^+^ T cell phenotype observed in this patient, and together support an increased differentiation of peripheral CD8^+^ T and NK cells in SLE patients with a recent or active IFN response.

**Figure 6.**
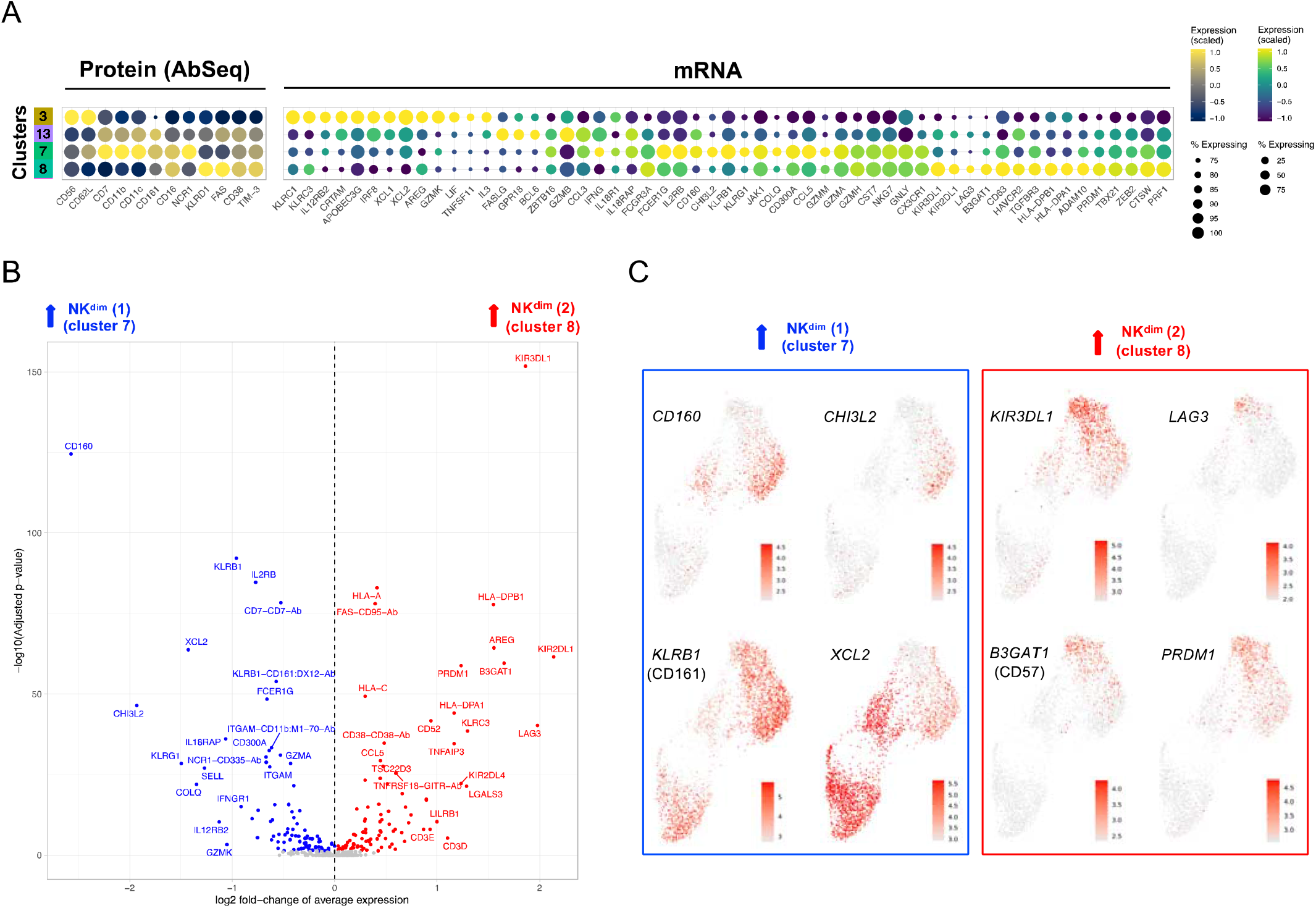
CD16^+^ NK^dim^ cells from the IFN^hi^ SLE patient display a highly differentiated phenotype. (**A**) Dot plot depicts the expression of select key differentially expressed markers within the identified NK cell clusters from the integrated analysis. The size of the dots represents the frequency of the marker in the respective cluster and the colour represents the relative expression level. Protein markers (AbSeq) are shown on the left section of the plots and mRNA markers on the right section. (**B**) Volcano plot depicts the differential expression between the two CD16^+^ NK^dim^ subsets (clusters 7 and 8) identified in the integrated analysis. (**C**) Feature plots depicting the expression of key differentially expressed markers between the CD16^+^ NK^dim^ subsets. Markers specifically upregulated in cluster 7 are highlighted in blue and markers specifically upregulated in cluster 8 are highlighted in red.

### Naïve T cells from the IFN^hi^ SLE patient display a signature of recent cytokine activation

Further supporting the systematic alterations induced by the inflammatory responses in patients with active disease, we identified a distinct signature of recent activation elicited by cytokine signalling within the CD4^+^ naïve T cells (cluster 1, as defined in Figure 5A) from the IFN^hi^ patient (Figure 7; underlying data – Dataset S7)^12^. This signature was consistent with the IFN^hi^-specific recently-activated naïve CD4^+^ T cell cluster identified in the resting dataset (cluster 12; Figure 2A, 2B), and was marked by the strong upregulation of several genes associated with regulation of cytokine signalling, including *SOCS2, CISH, BHLHE40, CXCR4, PRDM1, DPP4* and *MYB*, as well as the upregulation of IL-2RA at the protein level (Figure 7A, 7B). Although this cytokine-induced recent activation signature was more prominent within the naive CD4^+^ T cells, it could also be detected in both naïve Tregs (cluster 4; Figure 7C) and naïve CD8^+^ T cells (cluster 5; Figure 7D), albeit with smaller differences, suggesting that peripheral naïve T cells are sensitive biomarkers of recent cytokine activation. Importantly, this signature was consistent with the high plasma concentrations of IL-2 and sCD25 detected in this IFN^hi^ SLE patient (Table 1), indicating that an increased pro-inflammatory cytokine milieu in patients with active disease could be responsible for the observed transcriptional signature in naïve T cells. Together these findings support that gradual activation-induced alterations of the peripheral immune system are associated with the chronic immune activation in IFN^hi^ patients, which can be robustly characterised due to the increased functional resolution of the T and NK cells provided by the targeted multi-omics approach used in this study.

**Figure 7.**
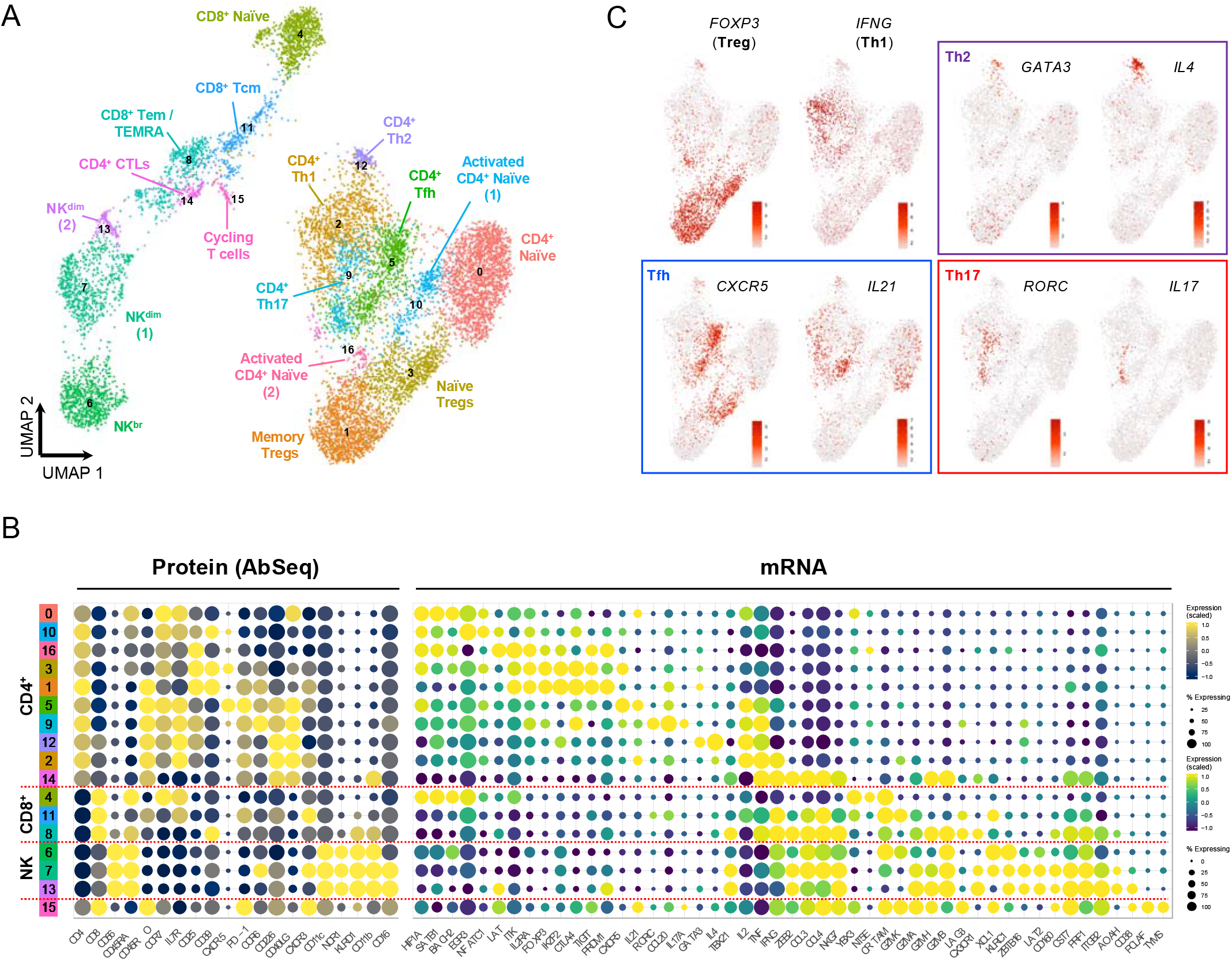
A subset of naïve CD4^+^ T cells from the IFN^hi^ SLE patient displays a cytokine-induced recent activation signature. (**A**) Volcano plot depicts the differential expression between the IFN^hi^ SLE patient and the two other donors in cells mapping to the naïve CD4^+^ T subset (cluster 1 of the integrated data). (**B**) Feature plots depict the expression of the cytokine-induced signalling gene *SOCS2* in the three donors profiled in this study. (**C**,**D**) Volcano plots depict the differential expression between the IFN^hi^ SLE patient and the two other donors in cells mapping to the CD4^+^ naïve Treg (cluster 4; **C**) and CD8^+^ naïve T cell (cluster 5; **D**) subsets, respectively.

## Discussion

The clinical management of SLE patients has been limited by the insufficient knowledge of the pathophysiological mechanisms that drive this complex disease and the limited available biomarkers to stratify patients. The recent advent of scRNA-seq technologies has promised to improve our understanding of the disease mechanisms, and shed new light on the specific contribution of the adaptive immune system in SLE^10^. Nevertheless, the current technical challenges and reduced sensitivity of scRNA-seq technologies based on transcriptome profiling only, where genes such as those encoding transcription factors that are expressed at low levels are difficult to detect, have not yet provided novel actionable insights to mechanistic-based patient stratification. The combination of protein quantification with traditional scRNA-seq technologies has been also implemented, and shown to significantly increase the power to discriminate immune populations^15,16^. However, these approaches have so far been mostly restricted to total PBMC samples, thereby reducing the number and diversity of the T and NK cells captured in these studies. To circumvent these limitations, in the present study, we have employed a targeted single-cell multi-omics approach providing both targeted mRNA and protein quantification, combined with pre-purification by flow sorting of the T and NK cell subsets, significantly increasing the number of relevant cells profiled and our confidence in assigning cell clusters to specific, and functionally relevant, sub-populations. This targeted approach – using a custom designed panel to assess the expression of 534 signature T and NK cell genes – not only provides increased sensitivity for the detection of the selected target transcripts, but also significantly decreases the costs of sequencing, thereby allowing to simultaneously quantify the expression of 51 surface expressed proteins. Here we show that this approach provides a high-resolution map of the peripheral T and NK cell populations in SLE in both resting and *in vitro* stimulated conditions. This increased resolution is particularly evident within the CD4^+^ T cell populations, where we are able to identify the different Th lineages in an informative way.

By applying this multiparametric deep phenotyping approach, we identified a subset of CD4^+^ CTLs showing hallmarks of highly differentiated T cells with a cytotoxic profile, which was specifically expanded in the IFN^hi^ SLE patient. This observation was consistent with our previous findings using this same technology on an independent IFN^hi^ SLE patient^11^, and further supports that the presence of a rare subset of CD4^+^ CTLs in blood, marked by the co-expression of surface markers usually expressed on cytotoxic CD8^+^ T cells, including CD57, CX3CR1 and KLRG1, is a feature of SLE patients with active disease. The description of a similar subset of differentiated CD4^+^ T cells with cytotoxic profile has been well established in humans^17^. Although the frequency of these cells, defined as perforin-expressing CD4^+^ T cells, is very low in healthy donors (usually <2% of CD4^+^ T cells in adults), they are significantly expanded and can reach frequencies of 5-20% of CD4^+^ T cells in patients with chronic viral infection^17^. More recently, scRNA-seq studies have further refined the characterisation of this CD4^+^ T cell subset, including CD57 expression, in the context of both viral infection^18^, anti-tumour responses^19^, and autoimmunity (primary Sjogren’s syndrome)^20^. In addition, a recent study has reported an increased frequency of CD4^+^ CTLs in super centenarians^21^. Taken together, these data strongly implicate CD4^+^ CTLs with antiviral response and chronic inflammation. Given that chronic IFN signalling is a central pathogenic factor in both primary Sjogren’s syndrome as well as in SLE, these findings could therefore provide a mechanistic rationale underlying the increased frequency of CD57^+^ CD4^+^ CTLs in blood of patients with active IFN responses. In support of this hypothesis, plasmacytoid dendritic cell activation, which is known to be one of the main sources of IFN production, has been shown to induce a pro-inflammatory cytokine profile in CD4^+^ T cells^22^. Furthermore, flow cytometric analysis of peripheral blood of SLE patients showed that a subset of memory CXCR3^+^PD-1^hi^ CD4^+^ T cells corresponded to these IFN-induced pro-inflammatory CD4^+^ T cells^22^. Of note, CXCR3^+^PD-1^hi^ CD4^+^ T cells were highly expanded in blood from SLE patients and showed a very similar cytotoxic profile to the CD4^+^ CTLs we describe in this study – which also express high levels of CXCR3 and PD-1. Given that CXCR3^+^PD-1^hi^ CD4^+^ T cells correspond to <5% of CD4^+^ T cells in healthy donors, it is plausible that an expansion of CD4^+^ CTLs is responsible for the observed increased frequency of CXCR3^+^PD-1^hi^ CD4^+^ T cells in SLE patients with active disease. This highlights the potential limitation of delineating immune subsets relying only on the expression of a small number of commonly expressed markers such as CXCR3 and PD-1, which could be preventing the identification of CD4^+^ CTLs and their specific contribution to the observed expansion of the more heterogeneous CXCR3^+^PD-1^hi^ CD4^+^ population in SLE. In the present study, we show that the expression of CD57 and CX3CR1 on the surface of CD4^+^ T cells could represent specific markers for the identification of CD4^+^ CTLs by flow cytometry. We therefore hypothesize that the frequency of CD57^+^CD4^+^ T cells in blood represents a novel cellular biomarker of recent or active IFN responses in patients with systemic autoimmunity, which could have informative clinical application for the monitoring of disease activity and represent a putative novel therapeutic target in SLE.

We note that in addition to the increased resolution of this targeted single-cell system, another key aspect of this study was the cell selection strategy, which allowed us to significantly increase the number of rare CD4^+^ T cell subsets. In particular, increasing the CD4^+^ CD127^low^ Teffs and Tregs populations, which are highly enriched for activated CD4^+^ T cells (such as the CD4^+^ CTLs), allowed us to identify rare subsets that could be otherwise difficult to robustly discern in more heterogeneous peripheral immune cell populations. Similar to the CD4^+^ CTLs, we also observed an expansion of terminally-differentiated CD8^+^ Tem/TEMRA cells with cytotoxic profile in the IFN^hi^ patient. An expansion of CD8^+^ CTLs has been previously reported in SLE patients with active disease at the single-cell level^10^. Although the expanded cytotoxic T cell cluster identified by Nehar-Belaid and colleagues displays hallmarks of CD8^+^ T cells, it is possible that it also encompasses the rarer subset of CD4^+^ CTLs, which as we show here display a very similar transcriptional profile to the CD8^+^ CTLs and may therefore not be discernible using whole-transcriptome single-cell technologies. Consistent with this putative pathogenic role of CD8^+^ CTLs in autoimmunity, a recent report has described a positive correlation between the frequency of a subset of CD57^+^ CD8^+^ T cells and the rate of C-peptide decline during the first two years after diagnosis in type 1 diabetic (T1D) patients^23^. In agreement with the CD8^+^ Tem/TEMRA subset identified in our study, the CD57^+^ CD8^+^ T cells described in recently diagnosed T1D patients display a classical cytotoxic profile marked by the expression of GZMB and CD16, and were shown to be enriched for autoreactive TCR clones, suggesting a pathogenic function in autoimmunity^23^. Our data add to a deeper phenotypic characterisation of these potentially pathogenic CD57^+^ CD8^+^ T cells, as well as their CD57^+^ CD4^+^ CTL counterparts, thereby providing a robust tool to characterise such disease-specific T cell subsets, and to identify markers that can be used to monitor their frequency using high-throughput methods, such as flow cytometry, in large numbers of autoimmune patients.

Further to the expansion of CD57^+^ CD4^+^ and CD8^+^ CTLs, we observed a similar increased expression of CD57 in the NK^dim^ cells of the IFN^hi^ SLE patient, reflecting an increased differentiation state in this patient. A reduction in the NK cell compartment has been reported in SLE using single-cell analysis^10^. This finding was consistent with previous reports using flow cytometry, which have also reported phenotypic alterations in NK^dim^ cells of SLE patients reflecting a similar increased activation profile^24,25^. The expression of CD57 in both CD8^+^ T cells and NK cells has been previously shown to be a marker of terminal differentiation, and the frequency of cytotoxic CD57^+^ cells has been shown to increase with age and to expand in a number of autoimmune diseases as well as in cancer^26^. Although the role of CD57 in CD4^+^ T cells is less well defined, it is plausible that it plays a similar role in all T cells, and is also associated with the terminal differentiation of a population of CD4^+^ CTLs. In addition, an expansion of CD57^+^ T and NK cells is also a feature of chronic viral infections, including human cytomegalovirus, human immunodeficiency virus, hepatitis C virus, and Epstein-Barr virus infections^26^. Given that the IFN gene expression signature in SLE is specifically induced by type I, but not type II, interferons, one hypothesis for the specific expansion of CD57^+^ cells in IFN^hi^ SLE patients is that the anti-viral type I IFN signalling pathway is directly linked with the increased differentiation of peripheral T and NK cells. These findings provide an additional cellular link between the chronic IFN signature detected in blood from approximately 50% of SLE patients and support the heightened activation profile in IFN^hi^ SLE patients.

A notable absence in this study was the identification of a distinctive IFN-induced gene expression signature in any of the defined immune subsets, which was the main source of biological variation driving the SLE-specific alterations characterised by single-cell analysis^10^. There are two main factors that could explain this observation: first is the targeted nature of the mRNA panel used in this study, which contains only a small number of IFN stimulated genes (ISGs). Nevertheless, we did measure the expression of several canonical ISGs, such as *OAS1, ISG15, IRF7, IFIH1, IFITM2* and *IFITM3*, and they were not significantly differentially expressed between the donors profiled in this study. Furthermore, given the widespread alterations induced by IFN stimulation, a significant number of the immune genes included in our panel should also be systematically regulated by an IFN response. A more plausible second explanation is that the expression of the ISGs is typically much less pronounced within T and NK cells compared to myeloid cell populations. Since only two SLE patients were profilled in this study, both of whom were recruited outside their regular clinic visits and had no evidence of flaring or active disease, it is likely that no discernible IFN signature could be identified at the time of sample collection in the peripheral T and NK cells of these patients. Instead, our findings suggest that more subtle, and long-lasting, immune alterations in the peripheral T and NK cell populations persist even during periods of clinical remission that are not associated with an active expression of ISGs. We believe that the application of this technology to a larger number of patients with a larger range of stages of disease activity will provide a clearer identification of the disease-specific IFN signature previously described in the T and NK cell populations using single-cell technologies^10^. Furthermore, applying this targeted approach to additional pathogenic immune populations typically associated with an IFN response, such as myeloid cells, may reveal increased heterogeneity of these populations and novel insights into the pathogenesis of SLE.

The major limitation of the current study is that only two patients and one healthy donor were profiled with this technology. The main aim of this study was to demonstrate the potential of applying a targeted approach - combined with a cell-type selection strategy - to dissect the heterogeneity of specific immune cell populations in more detail. By identifying a small set of patient samples with extreme phenotypic differences, we wanted to establish the applicability of this approach to identify more subtle disease-associated alterations in the adaptive immune compartment. This dataset therefore represents a proof-of-concept study and resource demonstrating the potential of this technology to immunophenotype the T and NK cell populations in SLE. These current findings therefore warrant further replication in larger, clinically well-characterised, cohorts; ideally in a longitudinal cohort to investigate subtle changes in the distribution of these immune populations over the course of the disease and the association with the underlying IFN response, as well as other clinical markers of disease activity. Given its targeted nature, this approach is highly scalable and could be applied to larger numbers of clinical samples and to other diseases, and has the power to robustly identify even rare immune subsets, whose signal may have been lost otherwise. These features are particularly well-suited to investigate the cellular mechanisms associated with response to therapy in large clinical trials. One such example is the use of ultra-low dose IL-2 therapy (three cycles of one million units of aldesleukin administered every other day for two weeks followed by a two-week interval), which has shown initial promise and a good safety profile in SLE^27^. More recently, these findings have been extended in a randomised clinical trial using the same dosing regimen^28^. However, one key conclusion from these studies is that response to IL-2 therapy is variable between patients, underscoring the clinical heterogeneity of SLE. In this study, He and colleagues reported that, compared to placebo, ultra-low dose IL-2 was very efficacious to induce clinical remission in patients with lupus nephritis and in patients with cutaneous rashes^28^. One potential confounding aspect in these trials is the diverse range of disease activity at the time of treatment, which can severely compromise the drug’s mechanism of action and consequently the attainment of primary clinical endpoints. For example, we have observed that the concentration of IL-2 in circulation and the frequency of Tregs were increased in SLE patients with increased disease activity^9^, and hypothesised that low-dose IL-2 treatment may be more effective for the maintenance of clinical remission rather than treating patients with acute or flaring disease - where additional exogenous IL-2 may be functionally redundant or proinflammatory. Indeed, the IFN^hi^ patient investigated here had high plasma IL-2 and sCD25 concentrations. These findings underscore the urgent need to better understand the mechanism of action of current drugs and to identify better biomarkers associated with clinical response and non-response to therapy. We note that an IL-2 mutein, AMG592 (efavaleukin alfa), is currently being investigated in SLE (registration number: NCT03451422). A follow-up phase 2b study (registration number: NCT04680637) will be initiated soon in a much larger cohort of 320 SLE patients and comparing its effects between patients with high IL-2 levels and those with normal physiological IL-2 concentrations will be of interest. In future trials it will also be important to investigate if ultra-low dose IL-2 efficacy is highest in SLE patients with certain disease manifestations such as nephritis and rashes, as reported recently by He et al^28^.

In conclusion, we have identified distinct alterations in the composition of the T and NK cell compartment in a patient with a discernible peripheral IFN signature, and high concentration of circulating IL-2 and sCD25. Most notably we identified an expansion of a subset of CD4^+^ CTLs and a concomitant increased proportion of CD8^+^ and NK cells with a terminally differentiated profile. Although an active IFN response could not be detected in these adaptive immune cells, these results suggest that additional subtle alterations in the cell composition could be more long-lasting than the prototypical IFN transcriptional signature and may represent more stable biomarkers of recent disease activity. More widely, this current study provides a framework for the application of a targeted single-cell multi-omics methodology to investigate the cellular mechanisms of human complex disorders as well as the mechanism of response to current therapies, including ultra-low dose IL-2. This information is currently largely lacking from most clinical studies and could provide critical insight for the design of future trials, including patient selection and treatment strategy, which are critical to increase the success rate of clinical trials in such clinically heterogeneous diseases as SLE.

## Data Availability

Data availability
Underlying data
Open Science Framework (OSF). Supporting material for 'Single-cell multi-omics analysis reveals IFN-driven alterations in T lymphocytes and Natural Killer cells in systemic lupus erythematosus'. https://doi.org/10.17605/OSF.IO/EDCTN.
This project contains the following underlying data:
- Dataset S1: Processed gene expression data matrixes (mRNA and AbSeq UMI counts per cell) and sample tag assignment per cell for the resting and stimulated datasets.
- Dataset S2: Differentially expressed genes in the resting cell clusters.
- Dataset S3: Differentially expressed genes in resting CD4+ CTLs (cluster 10 from the resting analysis) compared to remaining CD4+ T cell clusters.
- Dataset S4: Differentially expressed genes in the in vitro stimulated cell clusters.
- Dataset S5: Differentially expressed genes in the integrated resting and in vitro stimulated cell clusters.
- Dataset S6: Differentially expressed genes between the CD16+ NKdim cells in clusters 7 and 8 from the integrated analysis.
- Dataset S7: Differentially expressed genes in CD4+ naive T cells (cluster 1 from the integrated analysis) between the IFNhi SLE patient and the two IFNlow control donors.
Extended data
Open Science Framework (OSF). Supporting material for 'Single-cell multi-omics analysis reveals IFN-driven alterations in T lymphocytes and Natural Killer cells in systemic lupus erythematosus'. https://doi.org/10.17605/OSF.IO/EDCTN.
This project contains the following extended data:
- XLSX file containing targeted mRNA and protein (AbSeq) panels used in this study
Data are available under the terms of the Creative Commons Zero "No rights reserved" data waiver (CC0 1.0 Public domain dedication).

https://doi.org/10.17605/OSF.IO/EDCTN

## Abbreviations

CTL: cytotoxic T lymphocyte
FACS: fluorescence-activated cell sorting
IFN: type I interferon
IFN^hi^: SLE patient with a detectable type I interferon-inducible gene expression signature in blood
ISG: type I interferon-stimulated gene
NK: natural killer
PBMC: peripheral blood mononuclear cells
PMA: phorbol 12-myristate 13-acetate
scRNA-seq: single-cell RNA-sequencing
SLE: systemic lupus erythematosus
T1D: type 1 diabetes
Tcm: central memory T cell
Teff: effector T cell
Tem: effector memory T cell
TEMRA: terminally differentiated effector memory T cell
Tfh: follicular helper T cell
Treg: regulatory T cell
T1D: type 1 diabetes
UMAP: Uniform Manifold Approximation and Projection
UMI: unique molecular identifier.

## Data availability

### Underlying data

Open Science Framework (OSF). Supporting material for ‘Single-cell multi-omics analysis reveals IFN-driven alterations in T lymphocytes and Natural Killer cells in systemic lupus erythematosus’. https://doi.org/10.17605/OSF.IO/EDCTN^12^.

This project contains the following underlying data:

- Dataset S1: Processed gene expression data matrixes (mRNA and AbSeq UMI counts per cell) and sample tag assignment per cell for the resting and stimulated datasets.
- Dataset S2: Differentially expressed genes in the resting cell clusters.
- Dataset S3: Differentially expressed genes in resting CD4^+^ CTLs (cluster 10 from the resting analysis) compared to remaining CD4^+^ T cell clusters.
- Dataset S4: Differentially expressed genes in the *in vitro* stimulated cell clusters.
- Dataset S5: Differentially expressed genes in the integrated resting and *in vitro* stimulated cell clusters.
- Dataset S6: Differentially expressed genes between the CD16^+^ NK^dim^ cells in clusters 7 and 8 from the integrated analysis.
- Dataset S7: Differentially expressed genes in CD4^+^ naïve T cells (cluster 1 from the integrated analysis) between the IFN^hi^ SLE patient and the two IFN^low^ control donors.

### Extended data

Open Science Framework (OSF). Supporting material for ‘Single-cell multi-omics analysis reveals IFN-driven alterations in T lymphocytes and Natural Killer cells in systemic lupus erythematosus’. https://doi.org/10.17605/OSF.IO/EDCTN^12^.

This project contains the following extended data:

- XLSX file containing targeted mRNA and protein (AbSeq) panels used in this study

Data are available under the terms of the <u>Creative Commons Zero “ No rights reserved” data waiver</u> (CC0 1.0 Public domain dedication).

## Acknowledgments

We thank NIHR BioResource volunteers for their participation, and gratefully acknowledge NIHR BioResource centres, NHS Trusts, and staff for their contribution. We thank the National Institute for Health Research, NHS Blood and Transplant, and Health Data Research UK as part of the Digital Innovation Hub Programme. The views expressed are those of the authors and not necessarily those of the NHS, the NIHR or the Department of Health and Social Care.

